# Modeling COVID-19 vaccination strategies in LMICs considering uncertainty in viral evolution and immunity

**DOI:** 10.1101/2023.03.15.23287285

**Authors:** Daniel J. Klein, Luojun Yang, Cliff C. Kerr, Greer Fowler, Jamie A. Cohen

## Abstract

Vaccines against the SARS-CoV-2 virus were developed in record time, but their distribution has been highly unequal. With demand saturating in high-income countries, many low- and middle-income countries (LMIC) finally have an opportunity to acquire COVID-19 vaccines. But the pandemic has taken its toll, and a majority of LMIC populations have partial immunity to COVID-19 disease due primarily to viral infection. This existing immunity, combined with resource limitations, raises the question of how LMICs should prioritize COVID-19 vaccines relative to other competing health priorities. We modify an established computational model, Covasim, to address these questions in four diverse country-like settings under a variety of viral evolution, vaccine delivery, and novel immunity scenarios. Under continued Omicron-like viral evolution and mid-level immunity assumptions, results show that COVID-19 vaccines could avert up to 2 deaths per 1,000 doses if administered to high-risk (60+) populations as prime+boost or annual boosting campaigns. Similar immunization efforts reaching healthy children and adults would avert less than 0.1 deaths per 1,000 doses. Together, these modeling results can help to support normative guidelines and programmatic decision making towards objectively maximizing population health.

## 1 Introduction

Despite the rapid development of numerous vaccine products with high levels of protection against COVID-19 disease, the SARS-CoV-2 pandemic has been attributed with 15-18 million excess deaths globally by the end of 2021 (1; 2; 3), many of which occurred long after vaccines first became available. While vaccines are estimated to have averted 13.7-15.9 million deaths by the end of 2021 (4), vaccine distribution throughout the pandemic has been highly unequal and inequitable. Many high-income countries received vaccines early and in sufficient supply to fully vaccinate everyone, whereas a majority of low- and middle-income countries (LMICs) received few doses much later in the pandemic (5). This inequity persisted despite the best intentions and efforts of organizations like COVAX, which has distributed nearly 2 billion doses to LMICs (6).

Increasing vaccine availability, combined with decreasing demand in high-income settings, now raises the possibility of vaccinating populations that have thus far been left behind, including healthy children and adolescents in LMIC settings. With the recent waves of SARS-COV-2 spread, population immunity derived from infection has risen steeply in regions with low vaccine access, e.g., to 86.7% in Africa by December 2021 (7). While the safety and efficacy of COVID-19 vaccines has been well studied (8; 9; 10), the population value of vaccination has changed significantly as the pandemic has evolved (11). Taken together with reduced clinical severity associated with Omicron variants (12), these considerations prompt re-evaluation of the universal vaccination strategy adopted early during the pandemic, especially in low-resource settings where the burden of other diseases is high (13).

The efficacy and durability of COVID-19 vaccines depend on the trajectory of viral evolution and complex immune dynamics such as affinity maturation, imprinting, and short- and long-term immune memory (14; 15). While some studies observe that a third booster dose may only restore immunity to levels observed after the second dose (16), other studies suggest that additional boosters could increase peak naturalizing antibody levels with diminishing returns (17) or even continue to increase efficacy and immune breadth with each extra dose (18). In addition to the immune uncertainties, whether vaccination programs should focus on boosting high-risk populations or increase broader coverage by offering more priming series among children and adolescents or other groups without prior vaccine exposure remains an open question.

In this paper, we aim to evaluate the potential effectiveness and efficiency of COVID-19 vaccination programs through the end of 2025 in four archetypal LMIC settings. Our analysis utilizes an established agent-based model of COVID-19 dynamics, Covasim (19), to estimate the impact and efficiency of COVID-19 vaccines on averting health burden. In order to capture the effect of repeated immune events on affinity maturation, we adapt the immunological model in Covasim to incorporate increases in the breadth of antibody responses. We explore the effects of boosting frequency, priority subpopulations by age and vaccination history, and epidemiological settings on the long-term effectiveness and efficiency of COVID-19 vaccination programs. To the best of our knowledge, this is the first analysis to combine priming and boosting scenarios while also considering different pathways of viral evolution and addressing state-of-art immunological factors such as affinity maturation and diminishing returns on boosting.

## 2 Materials and methods

### 2.1 Base COVID-19 model

We used an adapted version of Covasim, an agent-based model of SARS-CoV-2 transmission and COVID-19 disease with detailed within-host dynamics and co-transmitting variants of concern. The base Covasim model is available as open-source code (20) and has been described elsewhere (19; 11). The model has been used throughout the pandemic to explore the impact of contact tracing in Seattle (21), explore school-based scenarios in the UK (22; 23; 24) and US (25), and support policy decisions in Australia (26), Vietnam (27), Poland (28), and elsewhere around the world.

Briefly, infections within the model can be asymptomatic, but if symptomatic have an age- and immunity-dependent risk of progressing to severe, critical, and death endpoints. Delays between stages are log-normally distributed with parameters that depend on the infecting variant. Infectivity is heterogeneous between individuals and elevated during the early stage of each infection.

Immunity in exposed or vaccinated individuals leverages established correlates of protection relating neutralizing antibody titers to efficacy against acquiring an infection, symptomatic disease, and severe disease (11) In the base model, each simulated individual has a single number representing their current antibody level that increases on each immune-inducing event (infection or vaccination) and wanes over time. However, with the emergence and co-circulation of variants of concern, it has become necessary to capture immune dynamics in greater detail.

### 2.2 Model adaptions for this analysis

We have modified the base model for this analysis to simulate separate neutralizing antibody titers for each possible source of immunity within each host. For example, an individual who had a wild-type infection early in the pandemic, received two doses of Pfizer vaccine, and later experienced an Omicron breakthrough infection will have separate antibody levels specific to each of these three sources (wild-type infection, Omicron infection, and Pfizer vaccine). Antibody levels and kinetics are source-specific with the first exposure to each source sampling a “priming” peak level from a log-normal distribution and subsequent source-specific exposures yielding a fold-increase “boosting” that acts multiplicatively on the previous peak. These channels act independently, representing separate B-cell populations.

Upon challenge with a specific viral variant, *j*, an “effective” neutralization correlate at time *t* is computed using a cross-neutralization matrix,

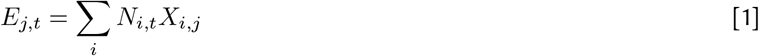

Here, *N*_*i,t*_ is the antibody level for variant *i* at time *t* within a host.

Values in the cross-neutralization matrix, *X*_*i,j*_ represent the fold-reduction in neutralization against variant *j* stemming from exposure to source *i*. These values are informed by live- and pseudovirus-neutralization studies assessing the antigenic distance between variants. The matrix is akin to a 2-dimensional antigenic map (29), with the added ability to capture asymmetries. After determining the effective neutralization for a particular challenge variant, *j*, established relationships (11) are used to calculate immune efficacy against infection, symptomatic disease, and severe disease.

Importantly, this analysis introduces two advanced immunological capabilities that have not been explored in other model-based analyses, to the best of our knowledge. The first new capability represents affinity maturation, the increase in immune breadth as a function of repeated exposures. Affinity maturation modifies the cross-immunity matrix, *X*, increasing cross-immunity (decreasing antigenic distance) between variants as a function of the number of vaccine doses an individual has received and if that individual has ever had a viral infection. Mathematically, the cross-immunity matrix is raised to a power, *θ*, which in turn is a function of exposure history, *h*,

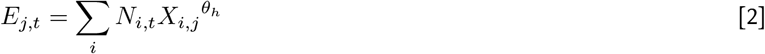

Increasing levels of exposure history result in lower powers that correspond to broader immune responses (0.6 *< θ <* 1 in the main analysis). For example, a cross-immune term between wild-type and Omicron could be as low as *X*_*wild,om*_ = 1*/*20 for an individual with no exposure history, but for *θ*_*h*_ = 0.6, this term would be modified to (1*/*20)^0.6^ ≈ 1*/*6, so a 6-fold reduction rather than a 20-fold reduction.

Due to high levels of scientific uncertainty in determining how affinity maturation, *θ*, depends on the exposure history, *h*, we consider two scenarios. Our main results are generated using a “mid-level” scenario in which boosters increase immune breadth (cross-neutralization) and also yield post-dose (peak) neutralization levels that increase with each dose, but with diminishing returns. For comparison, we also developed a “low-level” immune assumption as a pessimistic lower-bound on potential vaccine efficacy. The low-level scenario does not include any affinity maturation and boosting simply returns efficacy to the peak level observed after completion of the primary series. See the Supplemental Section on Immunological scenarios for additional information.

The second new capability represents diminishing returns on immunological benefits with increasing numbers of exposures. Covasim was initially designed during a period in which individuals would receive the primary series, and perhaps a single booster dose. The first dose results in “priming,” in which an initial peak neutralizing level is drawn from a source specific log-normal distribution. Subsequent doses capture “boosting” by multiplying the previous peak level by a constant factor, with no upper-bound imposed. While the constant factor assumption was reasonable in the early pandemic, it results in very high neutralization levels as individuals accumulate exposures over time. Instead, we specifically model diminishing returns; each dose has potential to yield peak neutralizing levels that are higher than achieved after the previous dose, but the fold increase can now decrease with increasing numbers of doses. Refer to the Supplemental Section on Immunological scenarios for additional details.

Infections within the model are traditionally established by infecting one or more naïve individuals to “seed” the outbreak in the modeled population. However, this approach requires significant human and computational time to calibrate and run the early period of the pandemic. Instead, for this analysis we imprint immunity from historical infections by selecting a specified number of people to infect with a specific variant on each historical day. We approximate the shape of each variant-specific historical wave as a Gaussian, and choose the amplitude, mean day, and variance for consistency with country-specific estimates of excess mortality from the WHO (1), IHME (2), and The Economist (3). The variant for each historical wave is informed by country-specific genomic surveillance (30). This process unfolds before the beginning of dynamic simulation, and thus there is no person-to-person transmission during this period. However, the full immune model capturing source-specific antibody priming, boosting, and waning is updated on a day-by-day basis. We also include immunity from historical vaccination during this period.

Dynamic simulation begins just prior to the Omicron wave in each country-like setting. During this period, infections are transmitted along an age- and setting -specific contact network with separate layers representing households, schools, workplaces, and community contacts (31). While individual person-to-person connections within each transmission layer can be can be dynamically modulated to represent physical distancing, lockdowns, school closures, and more, here we simply modulate overall transmissibility to roughly capture the impact of non-pharmaceutical interventions and behavior change during Omicron and subsequent historical waves. Transmissibility for the future period is determined by the most recent wave. As these are archetypal country-like settings, we simply selected transmissibility levels for broad consistency with estimates from IHME (32) and, importantly, the aforementioned excess mortality estimates.

### 2.3 Population contextualization

Considering recommendations described in the WHO COVID-19 Vaccine Prioritization Roadmap (33) depend on historical vaccine coverage, we selected archetypal settings representing a diversity of vaccine histories. This analysis includes Malawi-like, Kenya-like, South-Africa-like, and India-like settings representing low, middle, high, and very-high levels of historical vaccination, respectively. See Table 1 for additional details.

**Table 1.** Modeling assumptions for country-like settings as of December 31, 2023. For each archetypal setting, the table provides the population size and percent of population over the age of 60. The final two columns provide the percent of the full population that has completed the initial vaccine protocol and been boosted, respectively. Settings also differ in the timing of vaccine distribution, with India and South Africa earlier than Kenya, and Malawi increasing coverage more recently.

| Setting | Population | Population 60+ | Initial Protocol | Boosted |
| --- | --- | --- | --- | --- |
| Malawi | 18M | 4.1% | 22% | 0% |
| Kenya | 54M | 4.2% | 20% | 3% |
| South Africa | 59M | 8.5% | 33% | 7% |
| India | 1.4B | 10% | 68% | 14% |

Each country-like setting is intended to capture key elements of the pandemic in that region, but stops short of a full setting-specific calibration as would be needed for country-oriented policy making. Specifically, each country-like setting includes the age distribution of the population, virally-derived immunity from variant-specific historical waves of infections, and vaccine-derived immunity from time- and age-specific priming and boosting. We caution against deriving setting-specific policy from these archetypal representations because the simulations are at the national level and moreover were not informed by country experts regarding local epidemiology, interventions, care delivery, or other factors.

### 2.4 Viral evolution scenarios

A key determinant of future vaccine performance is viral evolution. The emergence of viral variants with diverse levels of transmissibility, severity, and immune escape has dominated the pandemic, especially with the emergence of Omicron in late 2021. Covasim does not simulate the mechanistic process of viral evolution endogenously. However, we exogenously introduce viral variants with complete flexibility including transmissibility, severity, immunogenicity, and cross-immunity from other variants and vaccines.

To capture uncertainty in viral evolution through the end of 2025, we consider two viral evolution scenarios, see Table 2. Each scenario captures viral evolution resulting in multiple waves of infections. In the “Continued Omicron” evolution scenario, we introduce a new Omicron sub-lineage variant on the first day of each year, beginning in 2023, with each new variant having the severity and transmissibility of Omicron and a two-fold escape from the previous variant. In the “New Cluster” evolution scenario, we simulate the emergence of a new viral variant on January 1, 2023, that is 10-fold away from Omicron as well as the wild-type cluster, including the ancestral variants like Alpha, Beta, Delta, and so on. Subsequent sub-lineages of this new cluster are introduced every six-months, each having a three-fold immune escape from the prior variant. Transmissibility and severity of variants in this cluster are assumed identical to that of Omicron. These scenarios are illustrated for the Malawi-like setting in Fig. 1.

**Table 2.** Viral evolution scenarios.

| Scenario | Cluster | Frequency | Immune Escape | Transmissibility | Severity |
| --- | --- | --- | --- | --- | --- |
| Continued Omicron | Omicron | New variant annually | 2x escape from previous | 1x | 1x |
| New Cluster | New | New variant every 6m | 10x escape initially, 3x thereafter | 1x | 1x |

Note: Transmissibility and severity are relative to Omicron BA.1.

**Fig. 1.**
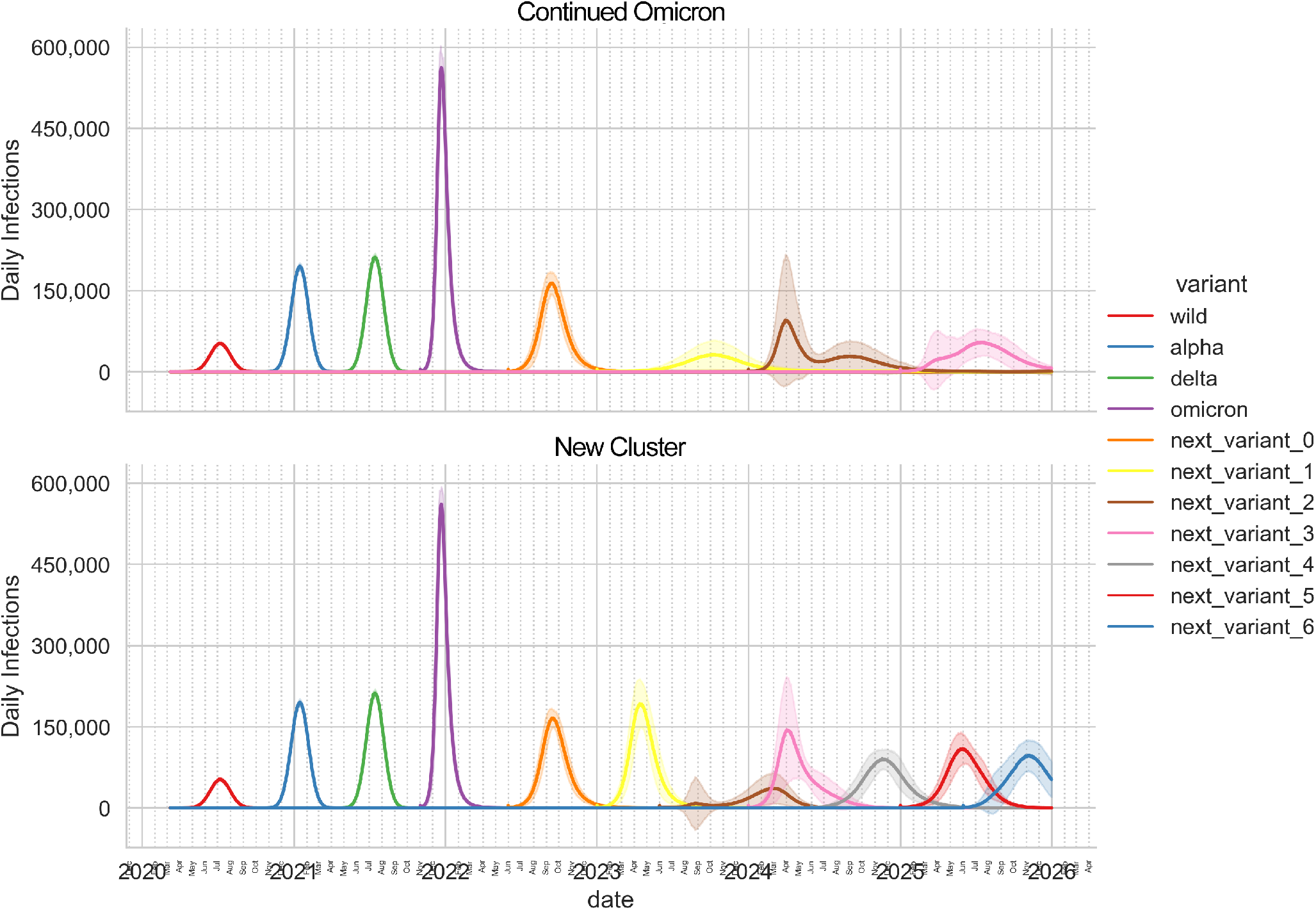
Historical calibration and viral evolution scenarios for the Malawi-like setting. The top and bottom panels show daily infections for the Continued Omicron and New Cluster evolution scenarios, respectively, in the status quo vaccine scenario that does not deliver vaccine in 2024 or 2025. This setting experienced historical waves from the wild, alpha, and delta variants, represented by red, blue, and green curves, respectively. Dynamic simulation begins with the “omicron” BA.1 variant and includes “next_variant_0,” which represents omicron BA.4 and BA.5. The evolution scenarios begin at the start of 2023. The Continued Omicron scenario includes one variant arriving on January 1 of each year. The New Cluster scenario includes a new cluster arriving on January 1, 2023 (yellow) followed by emergent sub-lineages within this cluster arriving every 6-months. Shaded regions represent two standard deviations above and below the mean (solid) of 50 model replicates.

### 2.5 Vaccine scenarios

The purpose of this analysis is to explore the potential benefits of a variety of future vaccine allocation scenarios in the archetypal country settings and against the two viral evolution scenarios described above. In each setting, the vaccination scenarios begin on January 1 of 2024 and continue for two years through December 31 of 2025. We explore priming and boosting vaccine scenarios prioritizing by age groupings ranging from children aged 6-months up to older adults aged 60+, see Table 3.

**Table 3.** Vaccine scenarios.

| Scenario | Timing | Age groups | Coverage |
| --- | --- | --- | --- |
| Status quo | No vaccination after Dec. 31, 2023 | N/A | N/A |
| Prime + boost | Priming at a constant rate over 2024, booster delivered 6m post-prime | 6m-12, 12-18, 18-60, 60+ | Increase coverage in the prioritized population group to 70% |
| Prime + 2x boost | As above + second booster 6m after the first booster |  |  |
| Annual booster | Boosting at a constant rate over 2024 and again one year later | 18-60, 60+ | 70% of the previously primed population |
| Bi-annual booster | Boosting at a constant rate over first 6mo of 2024, repeat every 6mo |  |  |

Considering the diversity of historical vaccine coverage levels in the represented settings, we begin with vaccine scenarios that reach populations that have not previously received vaccination. In addition to the priming regimen, these scenarios include one or two booster doses, delivered 6- and 12-months after completion of the primary series and reach populations aged 6-months to 12-years, 12-18 years, 18-60 years, and 60+. Within each country-age group, coverage is increased from the pre-scenario level to 70% linearly over the course of year 2024. We model a 10% dropout rate between completion of the primary series and first booster, as well as between the first and second booster doses for scenarios that include two boosters. Country-age priority groups that have already achieved 70% are excluded from this analysis and shown as “N/A” in the table.

Boosting scenarios complement the priming scenarios by delivering a dose annually or bi-annually to populations that have previously completed the primary series. We separately prioritize adults aged 18-60 years and older adults aged 60+ for this part of the analysis. In each routine boosting scenario, we assume that 70% of the previously primed population is reached. Vaccine is delivered at a constant rate over a one-year or six-month period, depending on the scenario. Subsequent doses are delivered to the prioritized individuals at exact 1-year or 6-month increments, depending on the scenario. Importantly, we assume strong correlation in doses in the sense that the subpopulation receiving additional vaccination is fixed at the individual level, representing a “compliant” core population.

All vaccine scenarios deliver a Pfizer-like mRNA vaccine and are compared against a Status Quo scenario in which no additional vaccine is delivered after December 31 of 2023.

### 2.6 Simulation methods and code availability

Dynamic simulation begins just prior to the Omicron wave in each country-like scenario and continues through December 31, 2025. Vaccine scenarios are compared against the Status Quo scenario during a two-year period starting January 1 of 2024, corresponding to the period over which additional vaccines are delivered as part of the scenarios.

The main analysis includes 4 country-like settings, 2 viral evolution scenarios, 2 immune variations, and 13 vaccine scenarios for a total of 104 configurations. Each configuration is replicated at least 250 times for a total of at least 52,000 simulations. Each of these replicates perturbs the seed of the random number generator to achieve results that are statistically significant from a stochastic model. Each country-like setting is represented by a statistical sample of the population using a total of 100,000 simulated individuals.

Updated code for the Covasim model is available from GitHub (20). The analysis repository is also available (34).

## 3 Results

Vaccine impact across country-like settings and vaccine scenarios is quantified using several key performance metrics including averted symptomatic infections, deaths, and years-of-life-lost (YLL). To aid in comparability to non-COVID vaccines, we also present results in terms of outcomes averted per fully vaccinated persons.

### 3.1 Impact of vaccination

All vaccine scenarios result in fewer deaths than the Status Quo scenario, with overall impact ranging from 2 to 40 deaths averted per 100,000 population over the two-year evaluation period, see Fig. 2. Across all country-like settings, approximately 20% more deaths are averted by vaccination for the New Cluster evolution scenario compared to the Continued Omicron evolution scenario. We also find that priming with two boosters is consistently more impactful by an average of 15% compared to priming with only one booster, and that routine boosting on a 6-monthly basis is more impactful than routine boosting on an annual basis, by a similar margin.

**Fig. 2.**
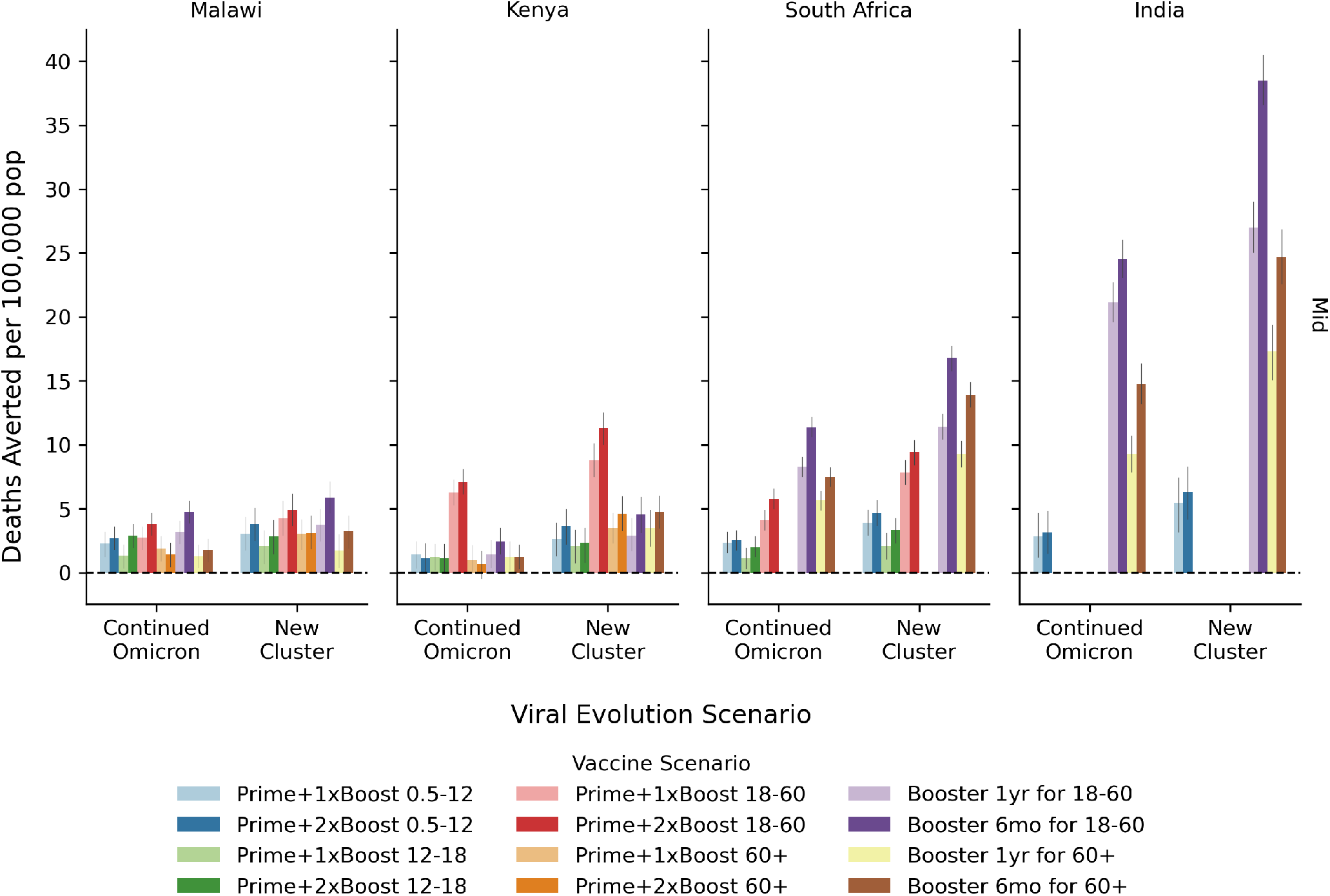
Vaccine impact. The overall impact of COVID-19 vaccination programs quantified in terms of the number of deaths averted over the two-year evaluation period per 100,000 people for the mid-level immunity assumptions. Within each country-like setting (columns) are two groups of bars corresponding to the viral evolution scenario. Bar color refers to the vaccine scenario, see the legend below the figure. The 95% confidence interval in the mean from 250+ model replicates is indicated by the gray vertical bar atop each colored bar.

We did not find large variation between country-like settings when considering deaths averted per 100,000 population. In the Malawi-like context, the greatest impact comes from boosting the adult (18-60 years old) population every 6 months with priming plus two boosters for the adult population a close second. In the Kenya-like setting, the scenario providing priming with one or two boosters to the adult population has the greatest impact. In the South Africa- and India-like settings, boosting the adult population has the greatest overall impact.

When considering impact quantified in terms of the number of infections averted per 100,000 population, we find that these vaccination scenarios avert 1,000 to 50,000 symptomatic infections per 100,000 population over the 2-year evaluation period with the mid-level immunity assumption, see Fig. S1 in Additional modeling results. Results considering symptomatic infections averted are more favorable to younger population priority groups, however the adult (18-60 year old) group achieves the largest impact in all settings. Across all scenarios, we find considerably lower vaccine impact with the low-level immunity assumption for infections averted (top figure panels) as well as deaths averted Fig. S2.

We have also computed the number of years of life lost that could be averted by these vaccine scenarios, see Fig. S3. With the mid-level immunity assumption, we find that these scenarios could avert between 25 and 700 YLLs over the two-year evaluation period per 100,000 people.

### 3.2 Efficiency of vaccination

Vaccine efficiency measures the number of outcomes averted per dose of vaccine delivered. Because the outcomes modeled in this analysis (deaths, symptomatic infections, and YLLs) are typically much smaller than the number of vaccine doses delivered, results will be presented per 1,000 doses delivered. The vaccine scenarios in consideration here deliver differing numbers of doses, ranging from two to four, and here we intentionally assess efficiency on a per-dose-delivered basis.

Results presented in Fig. 3 indicate that 1,000 vaccine doses can avert as many as two deaths in these scenarios. Across all country-like settings, we find a consistent pattern in which the scenarios prioritizing the 60+ population with either priming or boosting are by far the most efficient. In comparison, scenarios prioritizing children, adolescents, or even the general adult population are considerably less efficient.

**Fig. 3.**
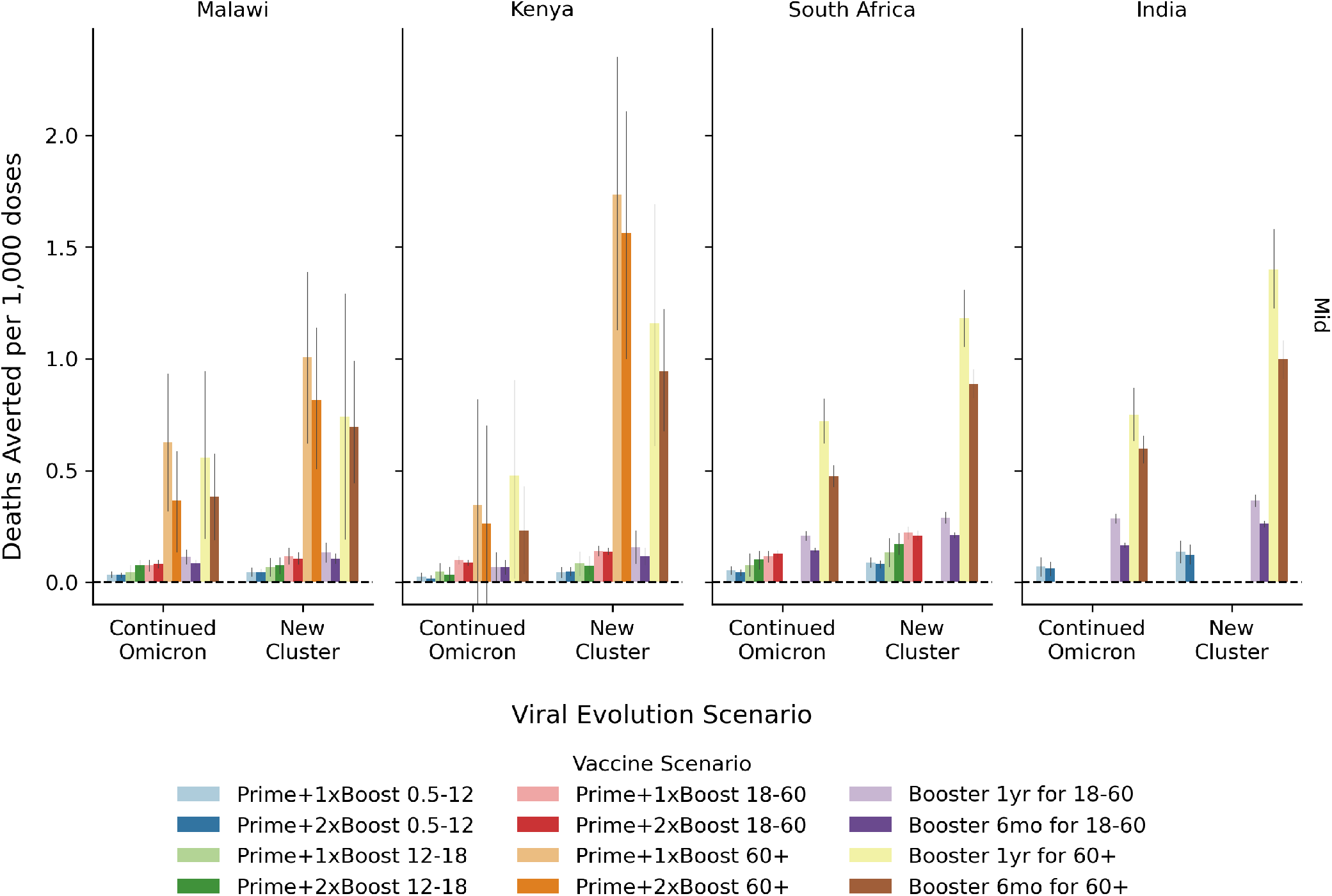
Vaccine efficiency. Efficiency of COVID-19 vaccination programs quantified in terms of the number of deaths averted per 1,000 doses over the two-year evaluation period per 100,000 people under the mid-level immunity assumptions. Within each country-like setting (columns) are two groups of bars corresponding to the viral evolution scenario. Bar color refers to the vaccine scenario, see the legend below the figure. The 95% confidence interval in the mean from 250+ model replicates is indicated by the gray vertical bar atop each colored bar.

Vaccination averts approximately 20% more deaths per 1,000 doses delivered when vaccinating against the New Cluster compared to a Continued Omicron viral evolution scenario, consistent with the impact results. In contrast with our impact results, we find that priming plus one booster is more efficient than priming plus two boosters and that routine boosters delivered annually are more efficient than routine booster delivered every 6-months, with the margin consistently about 20%.

Results for symptomatic infections averted per 1,000 doses are presented in Fig. S4. Here we find that 1,000 vaccine doses can avert between 50 and 1,250 symptomatic infections over the two-year evaluation period for the mid-level immunity assumptions, ignoring negative results that are due to stochastic variation and not statistically significant. Results are considerably lower for the low-level immunity assumptions. As compared to efficiency against death, these results are more uniform across scenarios with a slight preference for the general adult population in most country-like settings. Similar results showing years of life lost (YLLs) averted per 1,000 doses are available in Fig. S5, indicated that with mid-level immunity assumptions between 0 and 30 YLLs can be averted over per 1,000 doses. Results for the low-level immunity are close to zero and/or difficult to measure considering the stochastic variation in our model.

### 3.3 Efficiency per fully-vaccinated person

Because COVID vaccines are now competing for resources against other vaccine products, we present results quantified in terms of number of deaths averted per 1,000 fully vaccinated persons (FVPs), see Table 4 for results from the Continued Omicron viral evolution scenario. Each vaccine scenario defines a fully vaccinated person based upon a number of doses required, as specified by the “doses per FPV” number under each scenario listed in the table. Across all scenarios, deaths averted per 1,000 FVPs ranges from 0.06 to 2.38, depending primarily on the age priority group. Results reaching children 6m-12y and adolescents 12-18y are consistently low, below 0.3 deaths averted per 1,000 FVPs. Prioritizing the 60+ population for either the prime boost scenarios or the routine boosting scenarios achieves the greatest benefits with results generally in the range of 1-2 deaths averted per 1,000 FVPs.

**Table 4.** Deaths averted per 1,000 fully vaccinated persons under Continued Omicron evolution and realistic immunity assumptions.

|  | Age | Malawi | Kenya | South Africa | India |
| --- | --- | --- | --- | --- | --- |
| **Prime & Boost** |  |  |  |  |  |
| Prime + 1x Boost (3 doses per FVP) | 6m-12y | 0.10 [0.06, 0.14] | 0.07 [0.02, 0.12] | 0.12 [0.04, 0.19] | 0.21 [0.08, 0.33] |
|  | 12-18y | 0.13 [0.04, 0.22] | 0.14 [0.02, 0.25] | 0.20 [-0.01, 0.41] | N/A |
|  | 18-60y | 0.22 [0.15, 0.29] | 0.28 [0.24, 0.33] | 0.25 [0.15, 0.35] | N/A |
|  | 60+ | 1.82 [0.91, 2.72] | 1.00 [-0.44, 2.44] | N/A | N/A |
| Prime + 2x Boost (4 doses per FVP) | 6m-12y | 0.12 [0.08, 0.16] | 0.06 [0.00, 0.11] | 0.12 [0.05, 0.20] | 0.23 [0.11, 0.34] |
|  | 12-18y | 0.28 [0.19, 0.36] | 0.13 [0.00, 0.26] | 0.23 [0.00, 0.45] | N/A |
|  | 18-60y | 0.30 [0.23, 0.37] | 0.32 [0.28, 0.37] | 0.44 [0.35, 0.54] | N/A |
|  | 60+ | 1.35 [0.49, 2.22] | 0.98 [-0.64, 2.60] | N/A | N/A |
| **Routine boosting** |  |  |  |  |  |
| Booster 1/yr (2 doses per FVP) | 18-60 | 0.23 [0.16, 0.29] | 0.13 [0.02, 0.25] | 0.40 [0.34, 0.45] | 0.57 [0.53, 0.61] |
|  | 60+ | 1.11 [0.34, 1.89] | 0.96 [0.05, 1.87] | 1.38 [1.10, 1.65] | 1.50 [1.26, 1.73] |
| Booster 2/yr (4 doses per FVP) | 18-60 | 0.34 [0.28, 0.40] | 0.27 [0.15, 0.39] | 0.55 [0.49, 0.60] | 0.66 [0.62, 0.70] |
|  | 60+ | 1.54 [0.78, 2.30] | 0.92 [0.11, 1.73] | 1.75 [1.48, 2.01] | 2.38 [2.13, 2.63] |

Similar results including the the New Cluster evolution scenario as well as low-level immunity assumption are displayed in Fig. S6.

## 4 Discussion

It is likely that the SARS-CoV-2 virus will continue to evolve and cause infections and mortality well into the future. These modeling uniquely evaluate priming and boosting prioritization scenarios against uncertainty stemming from viral evolution and immunological factors like increasing breadth and diminishing returns on boosting.

Our modeling shows that the absolute impact of priming and boosting vaccine scenarios varies considerably across the four archetypal settings considered. These differences are driven by two factors. First is the number of doses distributed. The vaccine scenarios presented in this analysis are coverage-based. Country-like settings that have historically received little vaccine receive more doses to fill coverage gaps, and therefore yield more absolute impact. Second is the per-dose efficacy, which varies primarily as a function of the age of the vaccine recipient and secondarily as a function of prior immunity.

When considering dose efficiency, results consistently show that the highest returns come from reaching the populations at greatest risk for severe outcomes (the 60+ age group in this analysis). The relative inefficiency of scenarios prioritizing children and adolescents is striking in these results.

While not a perfect comparison due to methodological differences, our findings quantified in terms of deaths averted per 1,000 FVPs can be considered alongside a recent model-based analysis conducted by the Vaccine Impact Modeling Consortium (35). Toor et al. find that fully vaccinating 1,000 people could avert as many as 6.5, 7.7, or even 12 deaths for measles, hepatitis B, and human papillomavirus (HPV), respectively. Vaccines against yellow fever, Haemophilus influenzae (Hib), and pneumococcal disease all achieve over 2 deaths averted per 1,000 FVPs. In comparison, our results for the Continued Omicron evolution scenario only achieve values close to 2 deaths averted per 1,000 FVPs for the 60+ priority groupings. Results prioritizing children, adolescents, and even healthy adults are lower than would be expected from non-COVID vaccines.

Simulation results for the New Cluster scenario consistently yield vaccine impact and effectiveness results that are higher. However, this evolution scenario also has a lot more infections, severe outcomes, and deaths. The percentage of outcomes averted by vaccination is not dramatically different from the Continued Omicron evolution scenario.

Results for the “low-level” immune scenario are considerably more pessimistic than the “mid-level” scenario used in the main analysis. Additional data on the efficacy, durability, and breadth of increasing numbers of vaccine doses will be required to determine the relevance of this scenario.

The risk of death from an infection with SARS-CoV-2 increases exponentially with age. Our results include direct and indirect benefits of vaccination, but rapid waning and evolving variants ensure that the direct prevention benefits of vaccinating high-risk individuals outweighs indirect benefits from herd immunity.

The older age of deaths due to COVID-19 compared to many other vaccine-preventable diseases means that results quantified in terms of averted deaths will be more favorable for COVID vaccination programs compared to results quantified in terms of averted years of life lost and related health- or disability-adjusted life years lost. It is also worth consideration that COVID-19 vaccines are relatively expensive when compared to other vaccine products, particularly so for the mRNA-based vaccines. Beyond the unit cost for the vaccine itself, the scenarios represented in this manuscript could be quite complicated and ultimately expensive to deliver as programs specifically reaching these populations do not exist at scale in many LMICs.

### 4.1 Limitations

Like any modeling study, our results have many assumptions and limitations that could affect the results. First, while the population age structure closely resembles the national population in each country-like setting, the Covasim model does not simulate births or non-COVID deaths, dynamically age the population, nor modify contacts over time. Consequently, results may under-value vaccinating the youngest cohort. Second, vaccine benefits likely extend past the end date of the simulation, and therefore would be censored and under-estimated in this analysis. Third, vaccine is the only intervention considered in this analysis, which may artificially inflate the value of vaccine due to the lack of treatment, natural behavior change, or ongoing non-pharmaceutical policy interventions. Fourth, the model does not include key populations such as healthcare workers, pregnant women, nor immunocompromised populations; separate analyses will be required to inform vaccine prioritization decisions for these key populations. Fifth, model calibration to each country-like setting is a rough approximation at best. We used publicly available data on the age structure of each population, age-specific vaccination data, and circulating variants / case data, but did not work with country experts to refine each setting. For example, we do not mechanistically simulate the opening and closing of schools, and therefore may be over-estimating the prior exposure of children and adolescents, which in turn could apply a downward bias to our estimates of vaccine impact and efficiency in these populations.

Our analysis is also limited by data availability regarding vaccine efficacy. While the efficacy of the primary series was carefully studied in vaccine trials, recent data on the impact of third and even fourth boosters in diverse populations and against myriad variants has been more challenging to interpret. This modeling analysis therefore makes assumptions about the efficacy and durability of the fifth, sixth, and further doses. As the immune response to additional vaccine, especially in the context of ongoing transmission, is a key area of scientific uncertainty, we present a sensitivity analysis in the Supplementary Appendix.

## 5 Conclusion

Together, these modeling results suggest that vaccination against COVID-19 could be effective and efficient when prioritizing the highest-risk populations, those over 60+ in this analysis. Results are less favorable when considering averted years of life lost or other age priority groups. Healthy children and adolescents are a particularly inefficient use of vaccine in these results, suggesting that countries prioritize other vaccine programs before considering COVID-19 vaccines for these lower-risk populations. These results should be reassessed if/when new vaccines with increased durability and immunological breadth become available or if SARS-CoV-2 viral evolution takes an unexpected turn.

## Data Availability

All data produced are available online at
* https://github.com/institutefordiseasemodeling/covasim
* http://github.com/amath-idm/covid_vx_impact

http://github.com/amath-idm/covid_vx_impact

## Acknowledgments

We thank the many individuals who have contributed to the Covasim codebase or otherwise provided guidance and intellectual thought partnership along the way, specifically Mike Famulare, Robyn Stuart, Romesh Abeysuriya, Dina Mistry, Jasmina Panovska-Griffiths, Karen Makar, Mike Brison, Lynda Stuart, and Edward Wenger.

## 6 Supporting information

### 6.1 Immunological scenarios

Due to significant scientific uncertainty in immune function, we evaluated two immunological scenarios. These scenarios differ in the level of immune boosting and affinity maturation, ultimately manifesting in different trajectories of efficacy against infection, symptoms, and death related to ever-increasing numbers of vaccine doses. For the main analysis, we present an immunological scenario in which each dose results in a higher level of efficacy than modeled after the previous exposure. This is accomplished through the two mechanisms described in Section 2.2.

The first mechanism affects the neutralization level for a given vaccine source following a vaccine event. Unlike other models, Covasim tracks the peak titer following each event and for each immune source. Within each simulated individual, the neutralization level following a boosting event is determined by multiplying the previous peak level by a scalar,

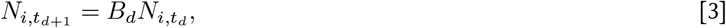

where *t*_*d*_ and *t*_*d*+1_ are the times of the neutralization peak levels following doses *d* and *d* + 1, respectively, and *B*_*d*_ is the boost factor as a function of dose. Boosting is not used for the first dose.

For the “mid-level” immune scenario, we assume the peak neutralization level increases four-fold on doses two and three, by two-fold on dose four, and returns to the previous peak level for five or more doses. As a sensitivity analysis, we consider a “low-level” immune scenario in which the peak neutralization level increases four-fold on doses two and returns to the previous peak level for three or more doses. See the “Boost factor” column in Table S1.

The second immune mechanism represents affinity maturation by the scalar *θ*, which varies according to previous immune history, *h* (see Eq. 2). Immune history is represented by previous exposure *ν* ∈ [0, 1] and the number of doses *d* ∈ [0, 1, 2, …] and individual has received. While this approach explicitly allows for hybrid immunity, it does not currently distinguish one previous exposure from two or more exposures.

For the “mid-level” immune scenario, we assume a degree of affinity maturation will accrue with increasing dose numbers and with viral exposure whereas for the “low-level” we disable affinity maturation. Vectors for *θ* displayed in Table S1 mathematically represent powers to which the cross immunity matrix is raised on an individual level, depending on their prior exposure and dose number, [0, 1, 2, 3, 4, 5+]. Lower numbers result in a greater degree of immune breadth.

**Table S1.** Immune scenario assumptions. Vectors correspond to dose numbers 1, 2, 3, 4, and 5+. Boosting is not used on the first dose, as indicated by *N/A*.

| Immune Scenario | Boost factor | Previous exposure? | Affinity maturation |
| --- | --- | --- | --- |
| Low-level | $B_d = [N/A, 4, 1, 1, 1]$ | Yes | $\theta = [1, 1, 1, 1, 1]$ |
| | | No | $\theta = [1, 1, 1, 1, 1]$ |
| Mid-level | $B_d = [N/A, 4, 4, 2, 1]$ | Yes | $\theta = [1, 1, 1, 0.7, 0.6]$ |
| | | No | $\theta = [1, 1, 0.7, 0.6, 0.6]$ |

### 6.2 Additional modeling results

Here we present supplemental model outputs to show how results vary across immune assumptions, viral evolution scenarios, and outcome metrics.

**Fig. S1.**
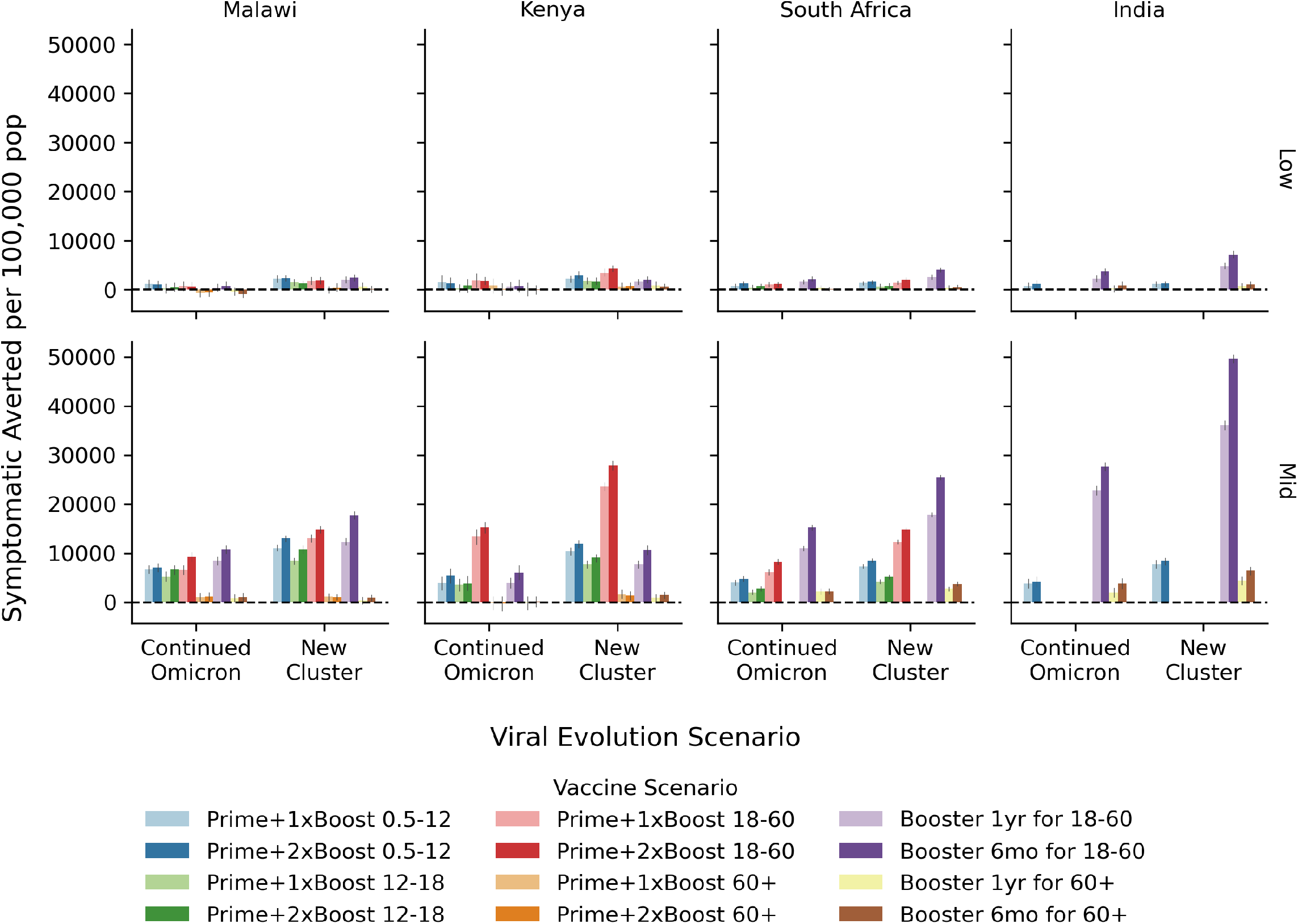
Vaccine impact: symptomatic infections averted. The overall impact of COVID-19 vaccination programs quantified in terms of the number of symptomatic infections averted over the two year evaluation period per 100,000 people for the mid-level immunity assumptions. Within each country-like setting (columns) are two groups of bars corresponding to the viral evolution scenario. Bar color refers to the vaccine scenario, see the legend below the figure. The 95% confidence interval in the mean from 250+ model replicates is indicated by the gray vertical bar atop each colored bar.

**Fig. S2.**
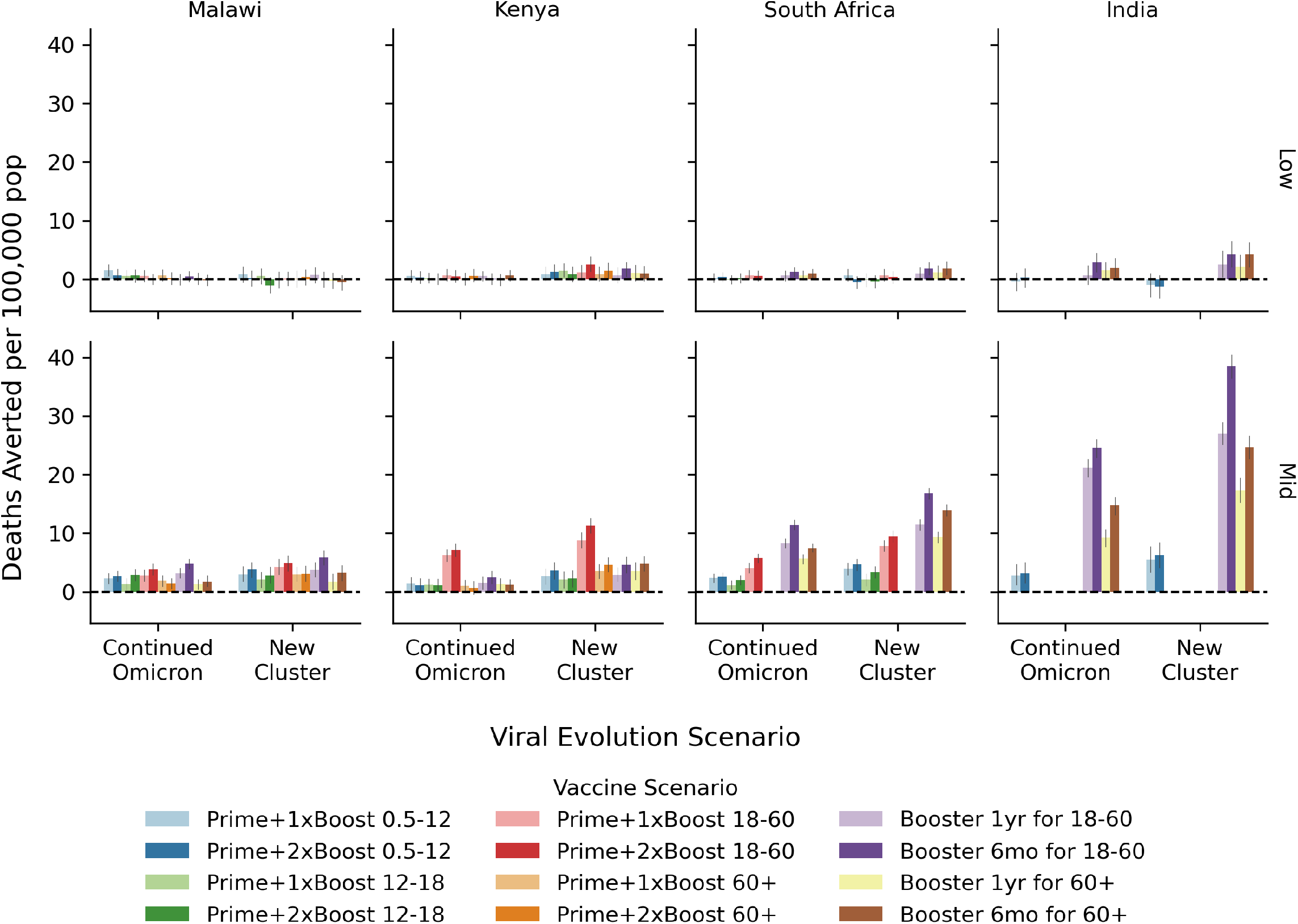
Vaccine impact: deaths averted. The overall impact of COVID-19 vaccination programs quantified in terms of the number of deathsaverted over the two year evaluation period per 100,000 people for the low- (top) and mid- (bottom) level immunity assumptions. Within each country-like setting (columns) are two groups of bars corresponding to the viral evolution scenario. Bar color refers to the vaccine scenario, see the legend below the figure. The 95% confidence interval in the mean from 250+ model replicates is indicated by the gray vertical bar atop each colored bar.

**Fig. S3.**
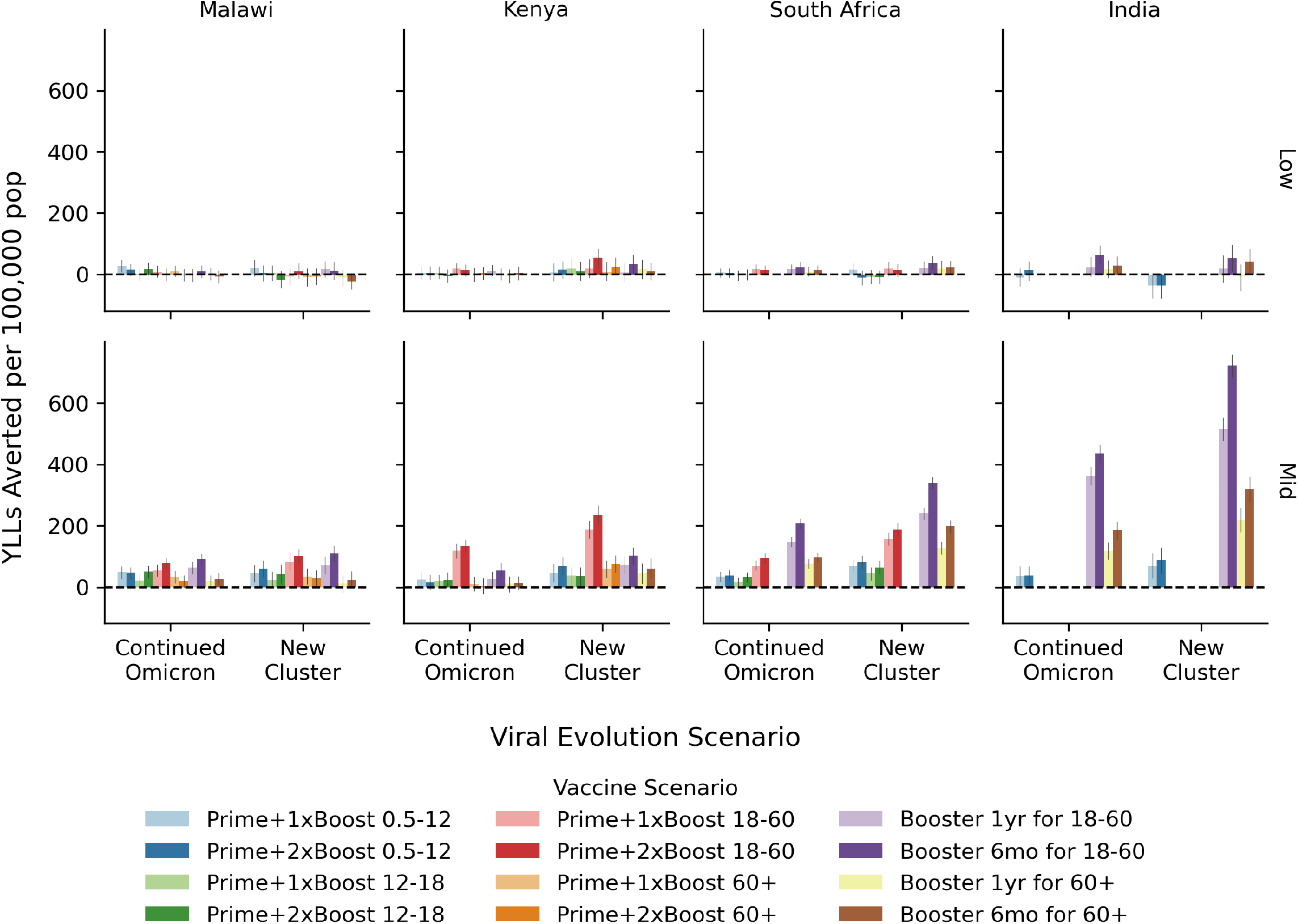
Vaccine impact: years of life lost averted. The overall impact of COVID-19 vaccination programs quantified in terms of the number of years of life lost that could be averted over the two year evaluation period per 100,000 people for the low- (top) and mid- (bottom) level immunity assumptions. Within each country-like setting (columns) are two groups of bars corresponding to the viral evolution scenario. Bar color refers to the vaccine scenario, see the legend below the figure. The 95% confidence interval in the mean from 250+ model replicates is indicated by the gray vertical bar atop each colored bar.

**Fig. S4.**
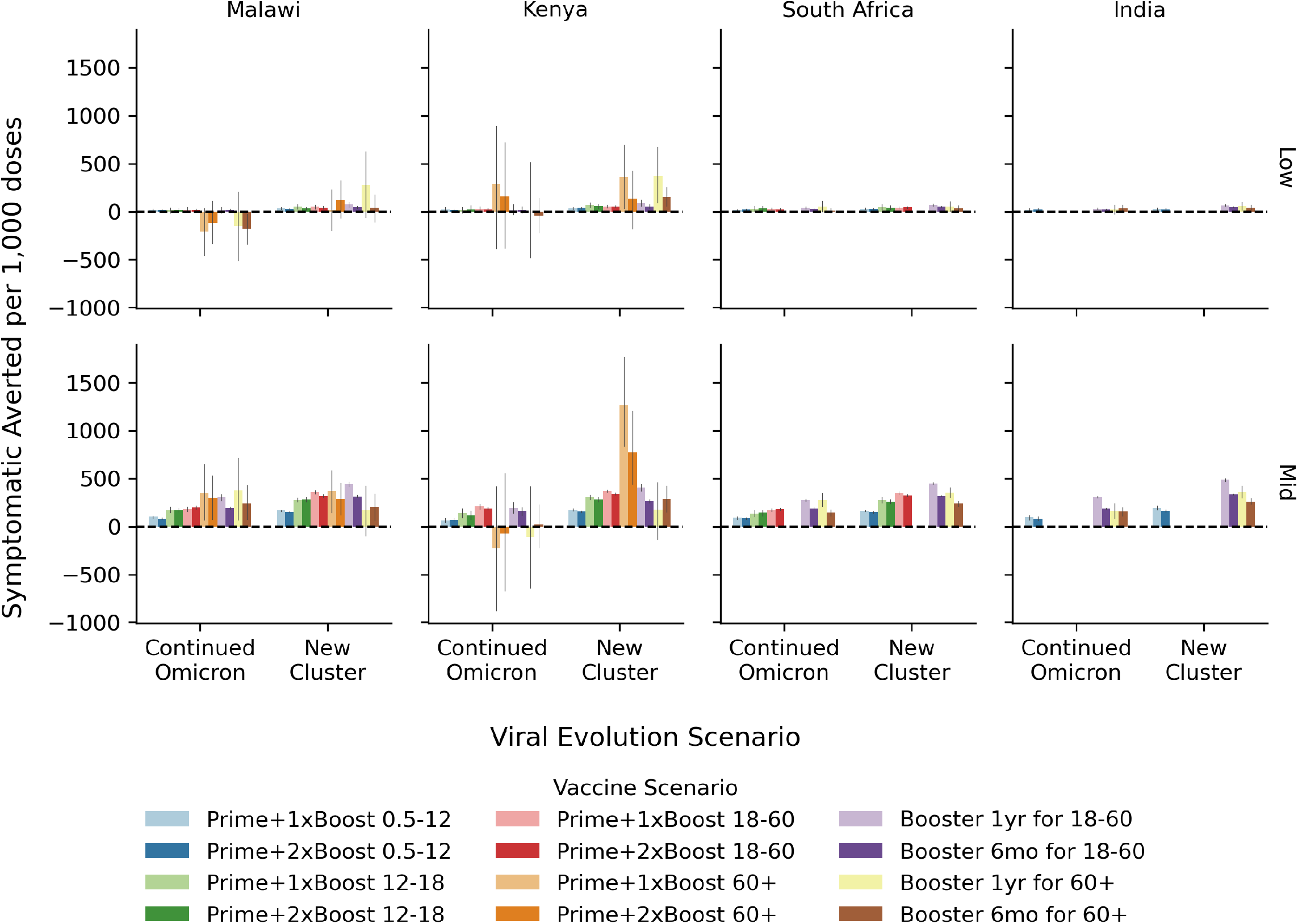
Vaccine efficiency: symptomatic infections averted. The efficiency of COVID-19 vaccination programs quantified in terms of the number of symptomatic infections averted over the two year evaluation period per 1,000 additional doses distributed for the low- (top) and mid- (bottom) level immunity assumptions. Within each country-like setting (columns) are two groups of bars corresponding to the viral evolution scenario. Bar color refers to the vaccine scenario, see the legend below the figure. The 95% confidence interval in the mean from 250+ model replicates is indicated by the gray vertical bar atop each colored bar.

**Fig. S5.**
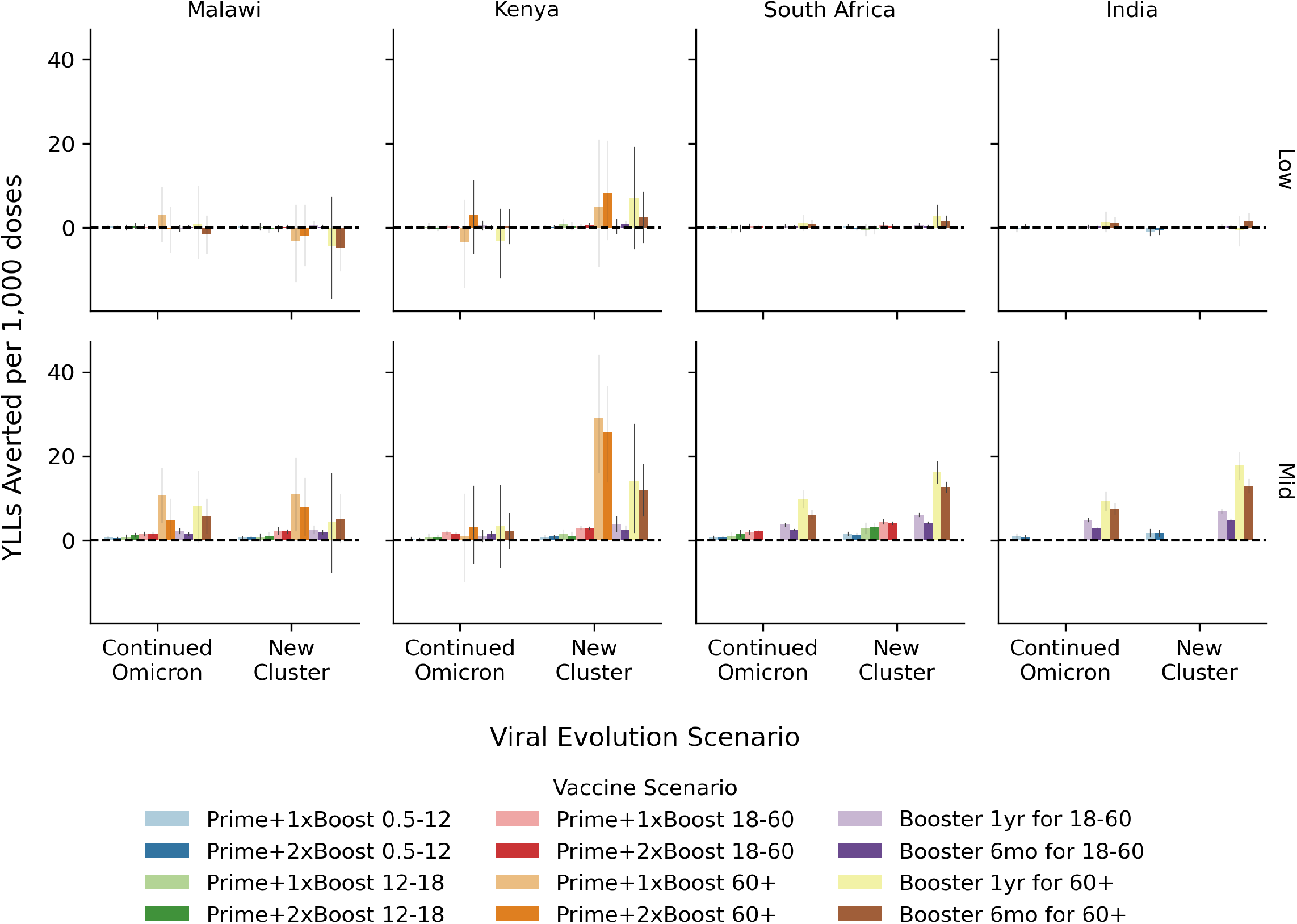
Vaccine efficiency: yeas of life lost averted. The efficiency of COVID-19 vaccination programs quantified in terms of the number of years of life lost averted over the two year evaluation period per 1,000 additional doses distributed for the low- (top) and mid- (bottom) level immunity assumptions. Within each country-like setting (columns) are two groups of bars corresponding to the viral evolution scenario. Bar color refers to the vaccine scenario, see the legend below the figure. The 95% confidence interval in the mean from 250+ model replicates is indicated by the gray vertical bar atop each colored bar.

**Fig. S6.**
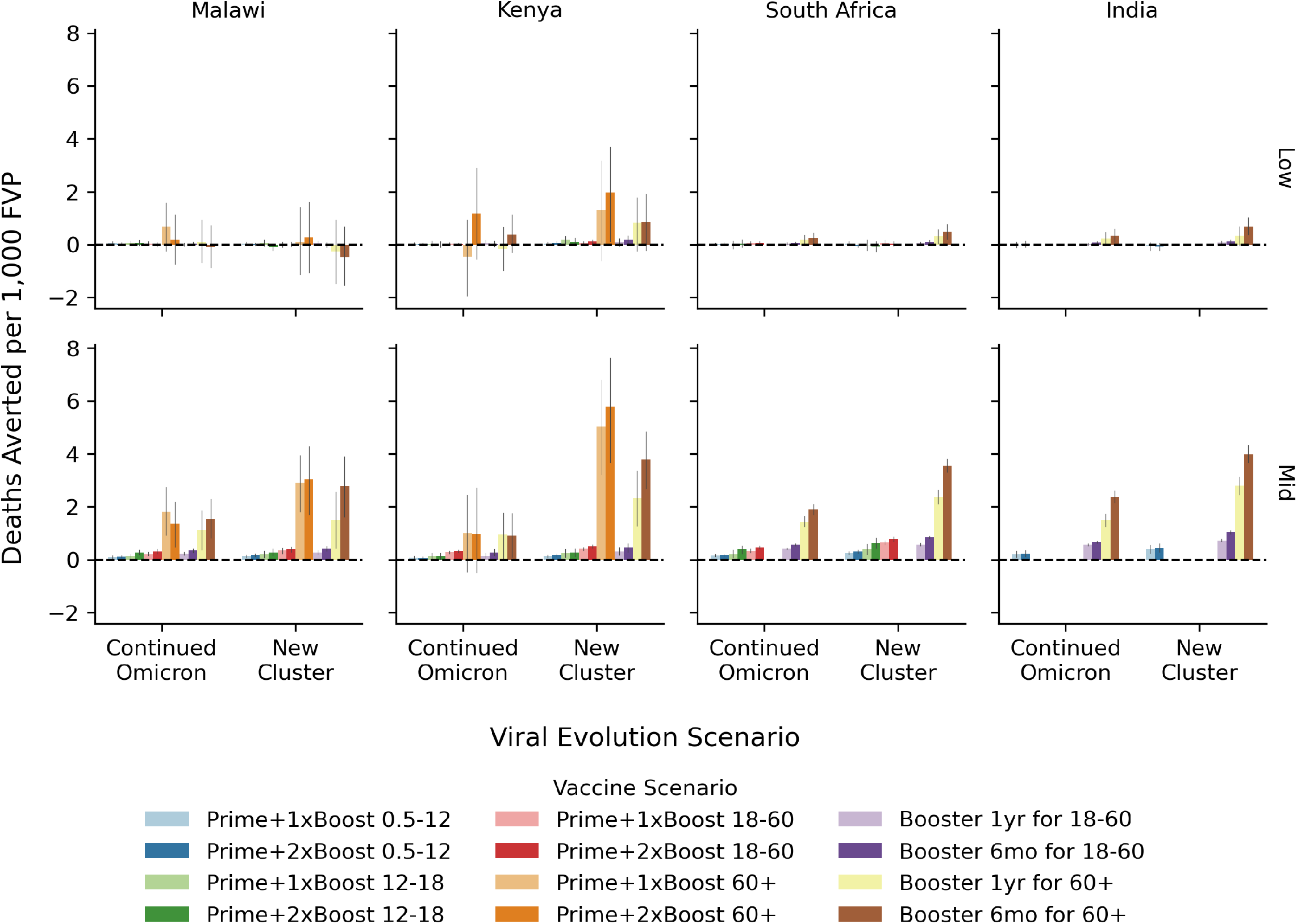
Vaccine efficiency (FVP): Deaths averted. The efficiency of COVID-19 vaccination programs quantified in terms of the number of deaths averted over the two year evaluation period per 1,000 fully vaccinated persons (FVPs) for the low- (top) and mid- (bottom) level immunity assumptions. Within each country-like setting (columns) are two groups of bars corresponding to the viral evolution scenario. Bar color refers to the vaccine scenario, see the legend below the figure. The 95% confidence interval in the mean from 250+ model replicates is indicated by the gray vertical bar atop each colored bar.

